# Systematic artifacts from Illumina two-color chemistry confound variant identification and actionability in clinical panels

**DOI:** 10.1101/2025.10.17.25338254

**Authors:** Hu Jin, Michail Andreopoulos, Vinayak V Viswanadham, Peter J Park

**Author notes:** These authors contributed equally: Hu Jin, Michail Andreopoulos.

## Abstract

Illumina short-read sequencing underpins clinical cancer genomics, with targeted panels widely used to detect actionable variants. Newer Illumina platforms employ a two-color chemistry to accelerate sequencing, but its impact on variant identification has not been systematically evaluated. Here we show that two-color platforms generate recurrent T>G artifacts in targeted panels at low variant allele fractions, predominantly occurring in specific trinucleotide contexts. These artifacts can produce spurious pathogenic variants in key cancer genes such as *TP53* and *KIT*, and inflate tumor mutational burden, a metric considered when assessing patient eligibility for immunotherapy. Accounting for such artifacts is therefore essential for accurate interpretation of clinical panel data.

## Main

Targeted panel sequencing is widely used in cancer clinics to identify somatic mutations that can inform diagnosis and treatment [1, 2]. Compared to whole-exome or whole-genome sequencing (WES/WGS), panels cover only a subset of the genome (∼50-500 cancer-related genes) but at much higher depth (∼500-2,000X), enabling sensitive detection of clinically actionable variants at lower cost. These variants can also be used to derive composite biomarkers such as tumor mutational burden (TMB) [3, 4], homologous recombination deficiency (HRD) scores [5, 6], and mismatch repair deficiency (MMRD) classifications [7, 8]. Despite their widespread use, reported mutations and aggregated metrics can vary substantially across panels due to differences in their covered regions, design (with or without matched controls), experimental protocols (tumor purity, sequencing depth), and bioinformatics pipelines (variant callers, parameters) [2, 9-11].

Illumina short-read sequencing remains the backbone of clinical genomics, with its sequencing-by-synthesis technology refined across successive platform generations. The earlier platforms— HiSeq and its lower-throughput version MiSeq—used a four-color chemistry, where each of the four DNA bases was tagged with a distinct fluorescent dye and imaged in a separate channel.

Newer platforms—NovaSeq and NextSeq—instead use a two-color chemistry, which halves the number of imaging cycles by encoding the four bases as unique combinations of presence and absence of two fluorescent signals (**Fig. 1a**). In this scheme, the absence of signal in both channels is interpreted as base G.

**Figure 1.**
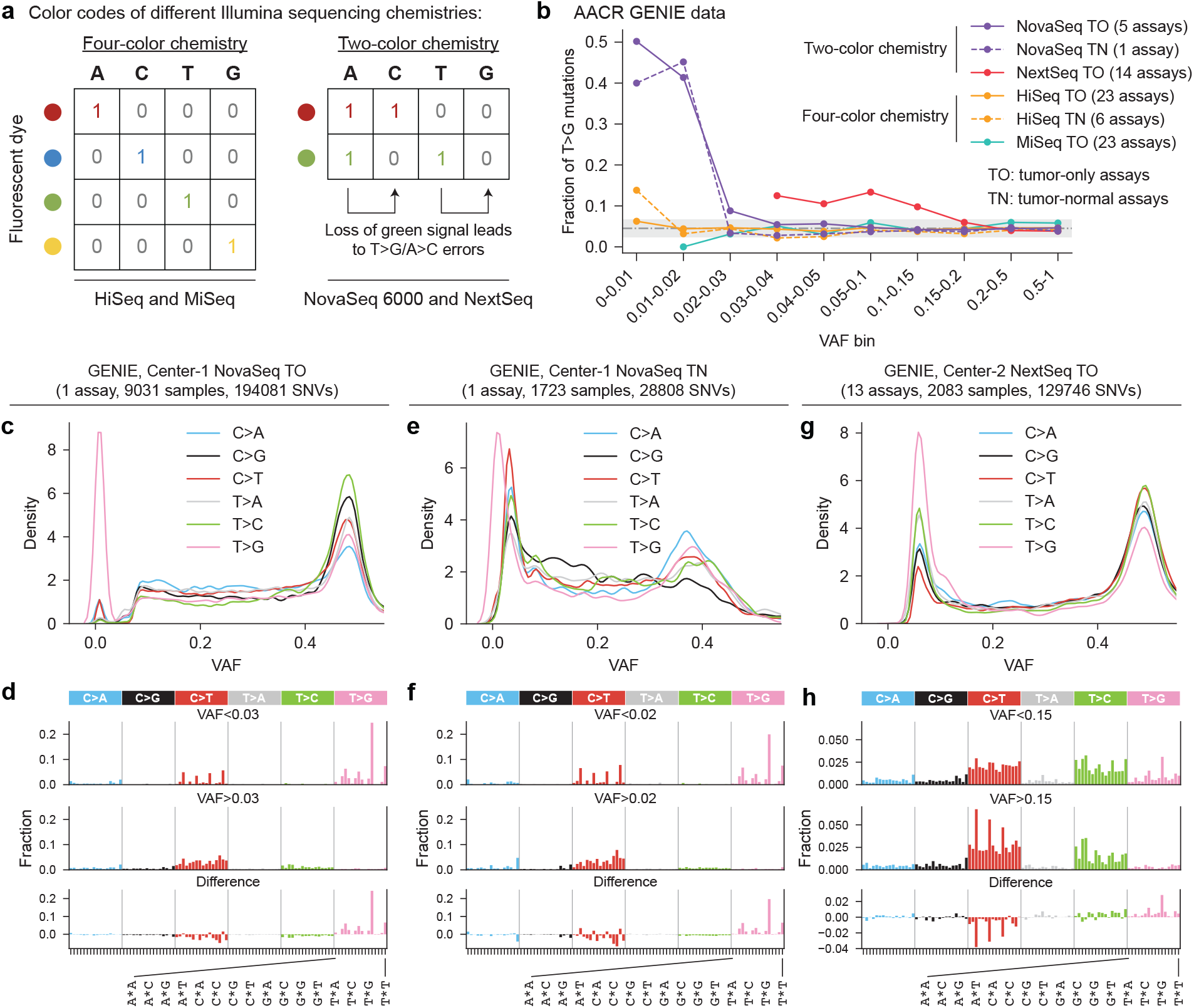
Targeted clinical panels are contaminated by recurrent T>G artifacts from Illumina two-color chemistry. **(a)** Schematic of color coding in four-color and two-color Illumina sequencing chemistries. **(b)** Fraction of T>G mutations across VAF bins in GENIE samples sequenced on different Illumina platforms. The gray dot-dash line indicates the mean T>G fraction in four-color assays, and the gray shading shows the mean ± three standard deviations. **(c)** Distribution of SNVs by substitution type across VAF ranges for sample set Center-1-NovaSeq-TO, showing enrichment of T>G mutations at low VAFs. **(d)** Mutational spectra of low-vs. high-VAF SNVs for sample set Center-1-NovaSeq-TO, together with their difference. **(e-f)** Same as (c-d) for Center-1-NovaSeq-TN samples. **(g-h)** Same as (c-d) for Center-2-NextSeq-TO samples.

However, because signal dropout can occur due to sporadic technical failures during sequencing, two-color chemistry is prone to systematic overcalling of G bases. This problem underlies the over-representation of poly-G sequences that require filtering [12]. More importantly, it can also lead to false positive somatic variant calls. A study comparing WGS data from three cancer cell lines sequenced with both chemistries reported an enrichment of T>G substitutions (equivalent to A>C) in two-color data [13]. Recently, we performed a comprehensive re-analysis of >10 publicly available WGS samples and further showed that these T>G artifacts can confound the detection of variants with low variant allele fractions (VAFs) in studies of mosaic mutations in non-tumor tissues [14].

Here, we report that such artifacts also occur widely in targeted clinical panels sequenced on two-color Illumina machines. In the AACR GENIE dataset [15]—one of the largest public cancer genomics datasets containing >200,000 panel samples—we identify recurrent T>G artifact mutations in key cancer genes such as *TP53* and *KIT*, mimicking pathogenic variants that would normally be considered clinically actionable. These artifacts can also inflate TMB estimates above the threshold (10 mut/Mb) used in FDA-approved tests for immune-checkpoint blockade (ICB) treatment [3, 4], potentially altering patient eligibility. With the phase-out of four-color platforms in 2024-2025 and two-color platforms becoming the standard, these artifacts must be accounted for when interpreting targeted panel data in cancer clinics.

## Results

### Low-VAF T>G mutations are enriched in panels sequenced on two-color Illumina platforms

Our analysis focused on somatic single-nucleotide variants (SNVs) in the ∼52,000 GENIE samples spanning 74 assays across 13 cancer centers, sequenced on Illumina machines and annotated with instrument model and VAF information. Different assays varied in panel designs, experimental protocols, or bioinformatics pipelines (**Supplementary Table**). After separating SNVs into six substitution types (C>A, C>G, C>T, T>A, T>C, and T>G), we calculated the proportion of T>G substitutions in different VAF ranges. Samples sequenced on four-color Illumina machines had an average of 4.5% T>G mutations, consistent across VAF bins (**Fig. 1b**). In contrast, samples sequenced on two-color machines were enriched for T>G mutations at low VAFs, regardless of whether the assay had tumor-only (TO) or tumor-normal (TN) designs (**Fig. 1b**). Using three standard deviations above the mean from four-color assays as a threshold, NovaSeq assays showed elevated T>G proportions at VAF<0.03 for TO assays and VAF<0.02 for TN assays, with up to 50% of SNVs being T>G at low VAFs—an 11-fold enrichment compared to four-color platforms. The T>G enrichment in NextSeq assays (all TO) was milder (up to 3-fold) but reached a higher VAF range (<0.15).

We next examined SNVs from each assay separately. A total of 13 assays from 4 cancer centers were sequenced on two-color machines and had sufficient data (≥1000 SNVs) for analysis. Among these, 11 assays from 2 centers showed T>G enrichment, including 2 NovaSeq assays from Center-1 and 9 NextSeq assays from Center-2 (**Supplementary Fig. 1a-b**). GENIE documentation indicated that the other 2 two-color assays (both NovaSeq) applied filters to exclude low-VAF (<0.03 or <0.05) mutations from reporting, which may have masked the T>G signal. By comparison, four-color assays generally displayed no such enrichment, even when examined separately (**Supplementary Fig. 1c-d**). Hereafter, we focus on the three sample sets with clear T>G enrichment: Center-1-NovaSeq-TO, Center-1-NovaSeq-TN, and Center-2-NextSeq-TO. Importantly, the apparent specificity to these two cancer centers primarily reflects the historical reliance of most GENIE contributors on older four-color machines and does not undermine the generalizability of the issue.

### Their mutational signatures are consistent with two-color sequencing artifacts

Mutational signature analysis is a powerful approach for inferring mutagenic processes from the characteristic sequence patterns of observed mutations [16-18]. Its application to cancer data has revealed a catalog of signatures corresponding to diverse endogenous and exogenous processes, as well as various sources of sequencing artifacts [19-21]. To validate that the enriched low-VAF T>G mutations in NovaSeq and NextSeq GENIE panels are artifacts of two-color chemistry, we compared their mutational signatures with the established two-color artifact signature—a predominant occurrence at NTG/NTT contexts—from our recent study (**Supplementary Fig. 2**) [14]. As expected, all three two-color sample sets exhibited a similar NTG/NTT signature for low-VAF T>G mutations but not at higher VAFs (**Fig. 2c-h**). In contrast, four-color samples did not show this signature at either low or higher VAFs (**Supplementary Fig. 3**). Moreover, the NTG/NTT signature was present in two-color but absent in four-color panels across nearly all tumor types, indicating that it reflects a sequencing artifact rather than a genuine subclonal mutational process (**Supplementary Fig. 4**).

**Figure 2.**
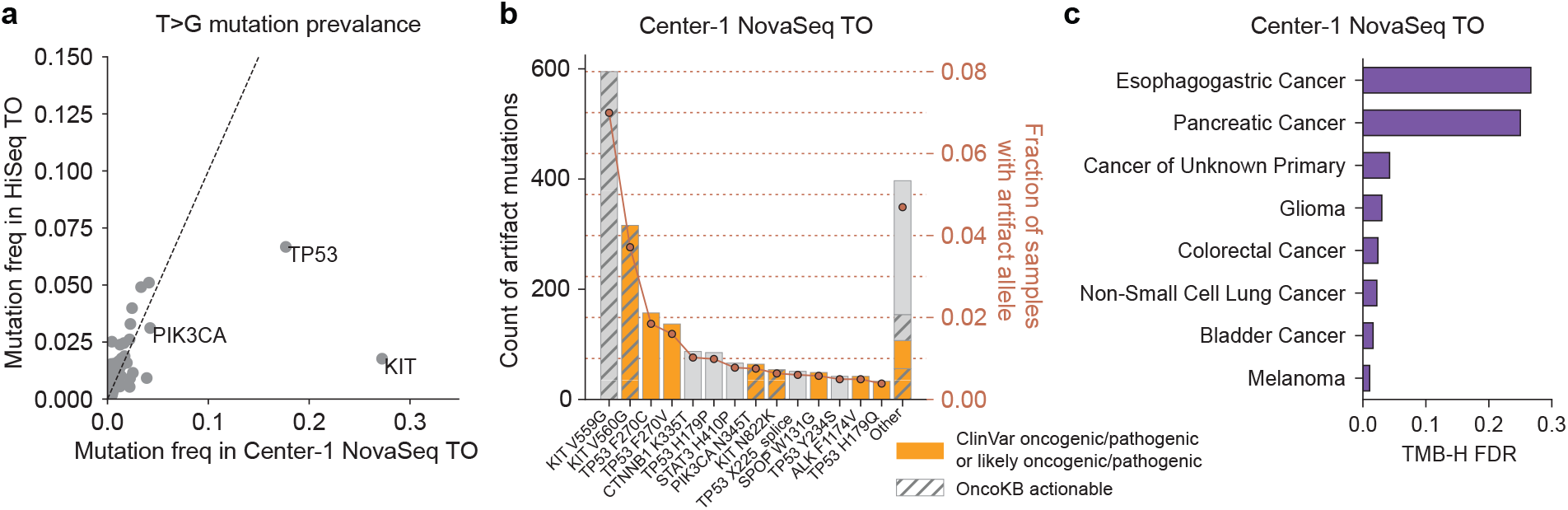
Two-color T>G artifacts confound clinical actionability of targeted panels. **(a)** Fraction of samples with a T>G substitution in each gene in Center-1-NovaSeq-TO compared with the four-color HiSeq baseline. Analyses were restricted to the same covered regions, and differences in tumor-type composition across assays were adjusted using a sampling scheme (Methods). **(b)** Counts of two-color T>G artifact alleles (bars) and the fraction of samples carrying each allele (dots) in Center-1-NovaSeq-TO. Analyses were restricted to T>G substitutions with VAFs below sample set-specific thresholds determined in Fig. 1b, and only non-silent, non-intronic alleles were included. Variants annotated as oncogenic/pathogenic or likely oncogenic/pathogenic in ClinVar are highlighted in orange. Clinically actionable variants according to OncoKB are indicated by grey hatching. **(c)** FDR of TMB-H samples due to two-color T>G artifacts across tumor types in Center-1-NovaSeq-TO. Only tumor types with ≥10 TMB-H samples before artifact removal were analyzed, and only tumor types with nonzero FDR were shown.

### T>G artifacts can generate false-positive pathogenic variants

To assess the clinical impact of these artifacts, we examined which variants and genes were most affected and whether they were annotated as pathogenic or clinically actionable in ClinVar and OncoKB [22-24]. At the gene level, several genes showed elevated frequencies of T>G mutations—irrespective of VAF—in two-color GENIE samples compared to the four-color baseline, after accounting for assay-specific target regions and cohort-specific tumor type compositions (Methods, **Fig. 2a, Supplementary Fig. 5**). We next restricted the analysis to T>G mutations likely to represent two-color artifacts, defined as those below sample set-specific VAF thresholds determined from **Fig. 1b**. Top affected genes and the percentages of samples with at least one T>G artifact in these genes are shown in **Supplementary Fig. 6**. In Center-1-NovaSeq-TO/TN, these genes included *KIT* (9.6%/7%), *TP53* (5.4%/4%), *CTNNB1* (1%/1%), and *PIK3CA* (1%/1%). In Center-2-NextSeq-TO, they included *NCOR2* (4%), *KMT2A* (1%), *MAP2K2* (3.5%), and *SOX10* (3.2%). When further restricted to non-silent, non-intronic T>G artifacts, 14.2%, 10.8%, and 21.3% of samples in Center-1-NovaSeq-TO, Center-1-NovaSeq-TN, and Center-2-NextSeq-TO, respectively, carried at least one such artifact mutation; 7.7%, 5.6%, and 0.6% contained at least one artifact annotated as oncogenic/pathogenic or likely oncogenic/pathogenic by ClinVar; and 9.8%, 6.8%, and 0.9% harbored at least one artifact annotated as clinically actionable in OncoKB. These percentages varied across tumor types (**Supplementary Fig. 7**). Several recurrent impacted variants were concerning, including *KIT* V559G/V560G, associated with sensitivity to KIT inhibitors such as imatinib in gastrointestinal stromal tumors and melanoma [25, 26]; *TP53* F270C/F270V, located within the DNA-binding domain of the protein and considered likely oncogenic [27, 28]; and *PIK3CA* N345T, linked to sensitivity to PI3K inhibitors (e.g., inavolisib) when combined with other drugs in some breast cancer subtypes [29] (**Fig. 2b, Supplementary Fig. 8**).

### T>G artifacts can inflate TMB estimates

TMB has been investigated as a predictive biomarker of clinical benefit from ICB. In 2020, FDA approved the PD-1 inhibitor pembrolizumab for adults and children with TMB-high (TMB-H) solid tumors, defined as those with TMB≥10 mut/Mb [3, 4]. We thus quantified the impact of two-color T>G artifacts on panel-based TMB estimates. An illustrative case is shown in **Supplementary Fig. 9a**, where ten T>G artifact mutations pushed the TMB estimate of a metastatic pancreatic cancer sample from 8.6 to 14.8 mut/Mb, resulting in a false-positive TMB-H classification.

Generalizing this example, we identified all false positive TMB-H calls in GENIE—that is, samples meeting the 10 mut/Mb threshold only because of likely two-color T>G artifacts, again determined using the sample set-specific VAF thresholds in **Fig. 1b**. TMB-H false discovery rates (FDR) were then calculated (**Fig. 2c, Supplementary Fig. 9b-c**). Tumor types with particularly high FDR included esophagogastric (27%) and pancreatic (25%) cancers in Center-1-NovaSeq-TO and head and neck cancer (21%) in Center-2-NextSeq-TO.

## Discussion

In summary, we identified a systematic T>G artifact in clinical panels caused by Illumina two-color chemistry and demonstrated that it can generate spurious actionable mutations in key cancer genes and inflated TMB estimates. Although these artifacts typically occurred at low VAFs, their significance should not be underestimated. First, VAF cutoffs for variant filtering are not yet standardized across clinical panels. For example, of the 16 clinical panel assays benchmarked by the Friends of Cancer Research TMB Harmonization Project, most assays did not apply VAF cutoffs but instead relied on minimum read counts supporting the variant allele, thereby allowing low-VAF variants to pass at high sequencing depth [10]. A survey of 50 clinical panels across multiple institutions reported that nearly half (24/50) applied VAF cutoffs ≤0.03 [30], which are too low to effectively filter out two-color artifacts that may be present in their data. Second, a uniform VAF threshold is not optimal, as it could obscure genuine variants present in subclones or in tumors with low purity. This issue is particularly relevant for NextSeq assays, where T>G artifacts occurred at VAFs up to 0.15 and thus overlapped with the range of true mutations, likely reflecting the higher overall error rate of NextSeq platforms described previously [31]. Clinically important driver mutations were also often reported even when they had lower VAFs. For example, the FDA-approved FoundationOne CDx assay lowers the VAF cutoff to 0.01 for base substitutions at cancer hotspots, whereas a higher cutoff of 0.05 is used otherwise [32]. Third, the challenge is magnified in cell-free DNA (cfDNA) panels, where true tumor variants occur at exceptionally low VAFs, potentially even below those of the two-color artifacts, as we previously observed for somatic mosaic mutations in deep WGS [14]. Given the increasing use of cfDNA panels for early cancer detection, residual disease monitoring, and TMB-based ICB selection, future work should evaluate the impact of two-color artifacts in cfDNA assays.

Our findings highlight the need for more effective strategies than a simple VAF cutoff to mitigate the impact of two-color artifacts. In clinical practice, when a low-VAF mutation occurs in the T>G (especially NTG/NTT) context and matches known artifact-prone sites (**Fig. 2b**), orthogonal validation should be considered, such as re-sequencing on a different platform. Curating a panel-of-normals composed of samples sequenced on two-color machines and incorporating it into a variant filtering step may help remove recurrent artifacts [13]. Ultimately, technological solutions at the sequencer level will be required. Illumina’s latest two-color sequencing chemistry, XLEAP-SBS, was introduced in 2023 for NovaSeq X and NextSeq 1000/2000 platforms. Whether this new chemistry mitigates two-color artifacts without introducing new sources of error remains unknown and requires further evaluation.

Finally, the two-color artifact we described here is by no means the only source of artifacts in clinical panels. For example, formalin fixation and paraffin embedding (FFPE), routinely used for processing patient material in the clinics, is well known to damage DNA and generate artifacts in somatic variant calls [33-35]. While analyzing GENIE data, we identified another distinct source of artifact T>C mutations specific to one assay, potentially caused by insufficient filtering of low-quality variants (**Supplementary Notes, Supplementary Fig. 10**). Systematic efforts are thus required to identify and catalog artifacts more thoroughly to ensure the fidelity of clinical panel sequencing.

## Methods

### Sample cohort

We analyzed data from AACR GENIE Release 17.0-public. Only assays sequenced on Illumina platforms with detailed instrument model information and samples with complete VAF data were included. VAF was calculated as “t_alt_count”/”t_depth”. Samples from Center-13 were excluded from all analyses except in the Supplementary Fig. 10 because of its assay-specific T>C artifacts. After filtering, the cohort consists of ∼800,000 SNVs from ∼52,000 samples, spanning 74 assays across 13 cancer centers. The cancer center and assay names were anonymized throughout the paper. Technical details of each assay can be found in Supplementary Table.

### Mutational signatures

Mutational signature analysis was performed by assigning each SNV into one of the 96 trinucleotide channels based on the substitution type and the 5’ and 3’ sequence context. Because different assays target different genomic regions with distinct background trinucleotide compositions, comparisons of mutational spectra across assays could be confounded. We therefore calculated assay-specific trinucleotide background frequencies and renormalized all mutational spectra to the whole-genome background. Specifically, let *x* denote the raw mutational spectrum, *W*_*panel*_ the trinucleotide background spectrum of a panel assay, and *W*_*WGS*_ the whole-genome background. Renormalization was performed as 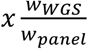, followed by L1 normalization of the resulting vector.

### Analysis of oncogenic/pathogenic and clinically actionable variants

To assess differences in the prevalence of T>G mutations across sample sets (Center-1-NovaSeq-TO, Center-1-NovaSeq-TN, and Center-2-NextSeq-TO), we compared each to a four-color HiSeq-TO baseline. For fair comparison, analyses were restricted to genomic regions shared across assays. Because of the large number of available HiSeq samples, we retained the six HiSeq assays with the broadest genomic coverage to ensure inclusion of key genes in both test and baseline cohorts. For each comparison, the number of patients and the tumor-type composition in the HiSeq baseline were matched to those of the test cohort by random subsampling from the larger HiSeq cohort. This sampling was repeated across multiple iterations, and the resulting error bars were negligible.

To evaluate oncogenicity/pathogenicity and clinical actionability, we leveraged the ClinVar and OncoKB databases [22-24]. Variants were classified as “oncogenic/pathogenic or likely oncogenic/pathogenic” if annotated as “Pathogenic”, “Pathogenic/Likely pathogenic”, or “Likely pathogenic” in the “ClinicalSignificance” field, or as “Oncogenic” or “Likely oncogenic” in the “Oncogenicity” field of the ClinVar file “variants_summary.txt”. Clinical actionability was determined using the OncoKB API, with a mutation classified as actionable if any level of evidence was present. These analyses were performed separately for each sample set (Center-1-NovaSeq-TO, Center-1-NovaSeq-TN, and Center-2-NextSeq-TO), restricting to T>G mutations likely to represent two-color artifacts (VAF<0.03 for Center-1-NovaSeq-TO, <0.02 for Center-1-NovaSeq-TN, and <0.15 for Center-2-NextSeq-TO).

### TMB estimates

TMB was calculated as the number of nonsynonymous mutations (SNVs and indels) per megabase of coding sequences (CDS) covered by each panel assay. Specifically, SNVs were included if annotated as “Missense_Mutation”, “Splice_Region”, “Nonsense_Mutation”, “Splice_Site”, “Nonstop_Mutation”, or “Translation_Start_Site” in the “Variant_Classification” field. Indels were included if annotated as “Frame_Shift_Del”, “Frame_Shift_Ins”, “In_Frame_Del”, “In_Frame_Ins”, “Splice_Site”, “Splice_Region”, “Nonsense_Mutation”, “Translation_Start_Site”, or “Nonstop_Mutation” in the “Variant_Classification” field. Variants annotated as oncogenic/pathogenic or likely oncogenic/pathogenic in ClinVar were excluded. CDS length was calculated by intersecting the covered genomic regions of each panel assay with CDS annotations of protein-coding genes from GENCODE (gencode.v47lift37.basic.annotation.gtf.gz).

## Data availability

AACR GENIE data were downloaded from the GENIE page on Synapse (https://www.synapse.org/#!Synapse:syn64501552). ClinVar data were downloaded from https://www.ncbi.nlm.nih.gov/clinvar/docs/ftp_primer/ftp.ncbi.nlm.nih.gov/pub/clinvar/tab_delimited/variant_summary.txt.gz.

## Code availability

Custom scripts for reproducing the analysis in this study are available on Zenodo (https://doi.org/10.5281/zenodo.17290056).

## Acknowledgements

This work was funded by grants from the National Institutes of Health (R01CA269805 to P.J.P.). The funders had no role in study design, data collection and analysis, decision to publish or preparation of the paper. The authors would like to acknowledge the American Association for Cancer Research and its financial and material support in the development of the AACR Project GENIE registry, as well as members of the consortium for their commitment to data sharing.

Interpretations are the responsibility of study authors.

