## Supplementary Notes and Figures for "Systematic artifacts from Illumina two-color chemistry confound variant identification and actionability in clinical panels"

#### Evidence of a separate T>C artifact associated with one GENIE assay

Although our analysis has focused on artifacts from Illumina two-color chemistry, this is not the only source of artifacts in clinical panels. For example, formalin fixation and paraffin embedding (FFPE), routinely used for processing patient material in the clinics, is well known to damage DNA and introduce artifacts in somatic variant calls [1, 2]. Here we describe another potential artifact—consisting of T>C mutations—observed in GENIE data. One assay in particular—Center-13-Assay-1 (sequenced on MiSeq)—displayed a much-elevated proportion of T>C mutations at low VAF (<0.1) compared to other HiSeq/MiSeq assays (**Supplementary Fig. 10a**). Mutational signature analysis revealed that these low-VAF T>C mutations predominantly occurred in the NTG/NTT contexts (**Supplementary Fig. 10b-c**), closely resembling an artifact signature reported by Guo *et al.* [2] in their reanalysis of the panel data from Prentice *et al.* [3] (Pearson $r$=0.66, $p$<0.005; **Supplementary Fig. 10d**). Among all HiSeq/MiSeq assays, low-VAF T>C mutations in Center-13-Assay-1 showed the strongest similarity to this T>C artifact signature (**Supplementary Fig. 10e**). In Guo *et al.*, the authors relaxed the variant filters originally applied in Prentice *et al.*, allowing additional low-quality mutation calls to be included. This revealed two artifact signatures enriched among these calls: a C>T signature attributed to FFPE and a T>C signature of unknown origin, present in both FFPE and non-FFPE samples. We hypothesized that the excess low-VAF T>C mutations in Center-13-Assay-1 similarly reflect insufficient filtering of low-quality variant calls analogous to Guo *et al.*, where variant filters were deliberately relaxed. Because all Center-13-Assay-1 specimens were FFPE-preserved, this hypothesis further predicted that FFPE-associated artifacts should also be present in this assay. Consistent with this, low-VAF C>T mutations in Center-13-Assay-1 closely followed the FFPE artifact signature from Guo *et al.* (Pearson $r$=0.94, $p<10^{-7}$), more so than in any other HiSeq/MiSeq assay (**Supplementary Fig. 10f-g**). Finally, as with the two-color artifacts, these T>C artifacts can confound clinical interpretation by introducing spurious mutations in key cancer genes (**Supplementary Fig. 10h-i**).

### Supplementary Figures

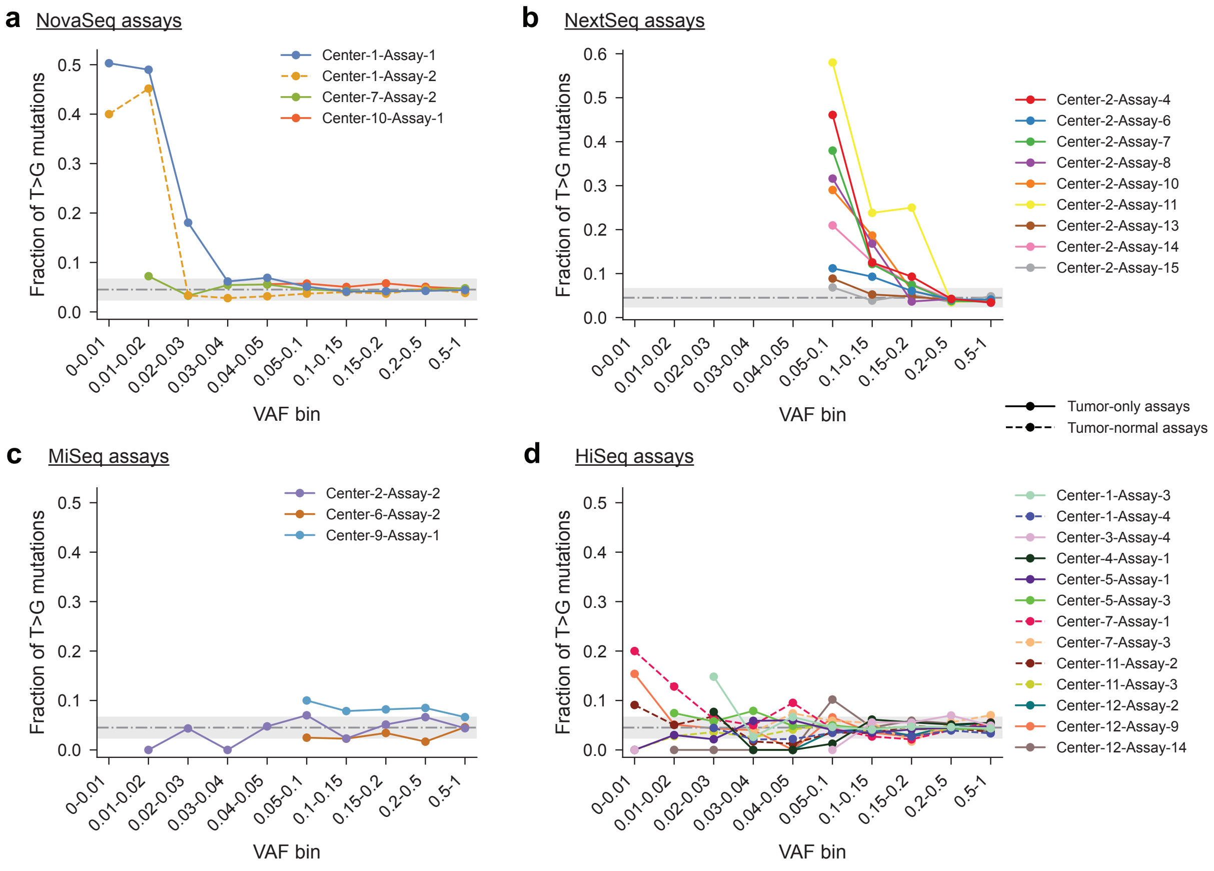


***Supplementary Figure 1. Fraction of T>G mutations across VAF bins separately for each assay sequenced on NovaSeq (a), NextSeq (b), MiSeq (c), and HiSeq (d)****. Only assays with at least 1,000 SNVs were included.*

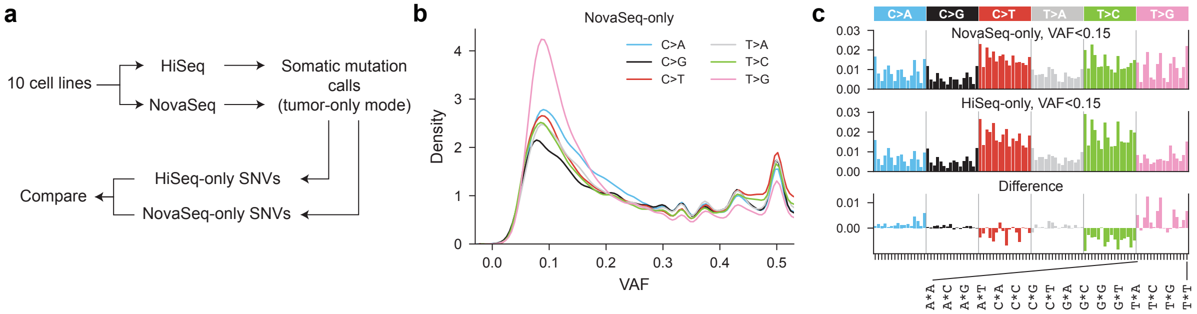


***Supplementary Figure 2. Discovery of the two-color artifact signature in our recent study [4]. (a)*** *Study design.* ***(b)*** *Distribution of SNVs by substitution type across VAF ranges, showing enrichment of T>G mutations at low VAF.* ***(c)*** *Mutational spectra of low-VAF SNVs uniquely detected in NovaSeq ("NovaSeq-only") and HiSeq ("HiSeq-only") data, together with their difference.*

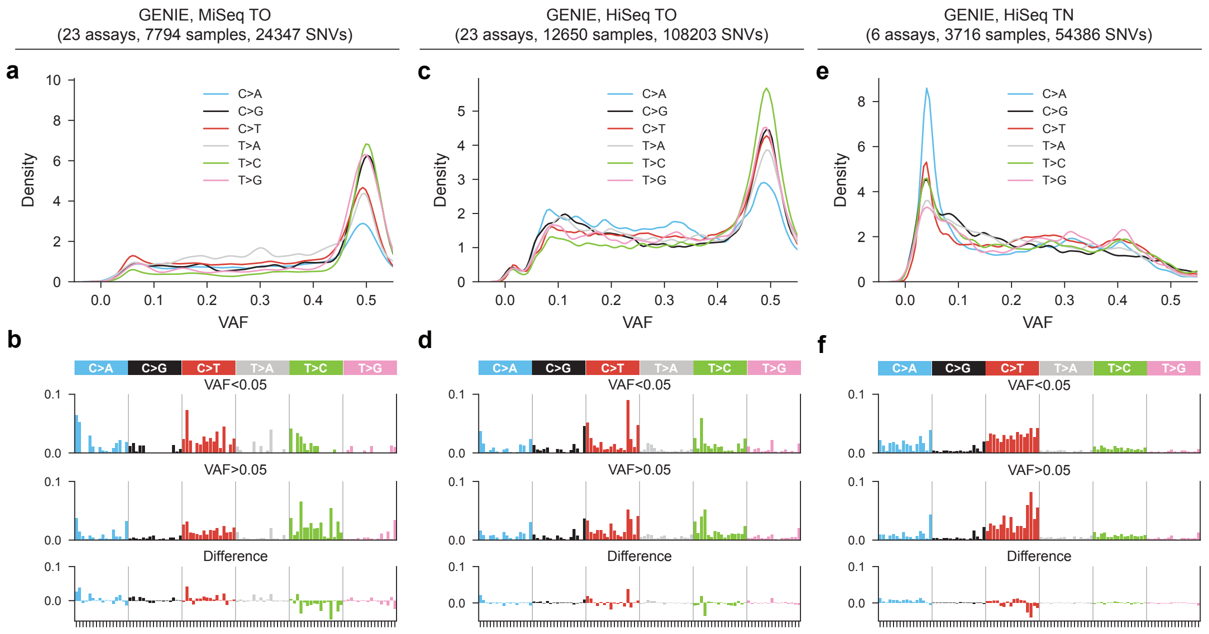


***Supplementary Figure 3. Mutational signature analysis of SNVs in GENIE samples sequenced on four-color assays. (a-b)*** *Same as Fig. 1c-d for MiSeq TO samples.* ***(c-d)*** *Same as Fig. 1c-d for HiSeq TO samples.* ***(e-f)*** *Same as Fig. 1c-d for HiSeq TN samples.*

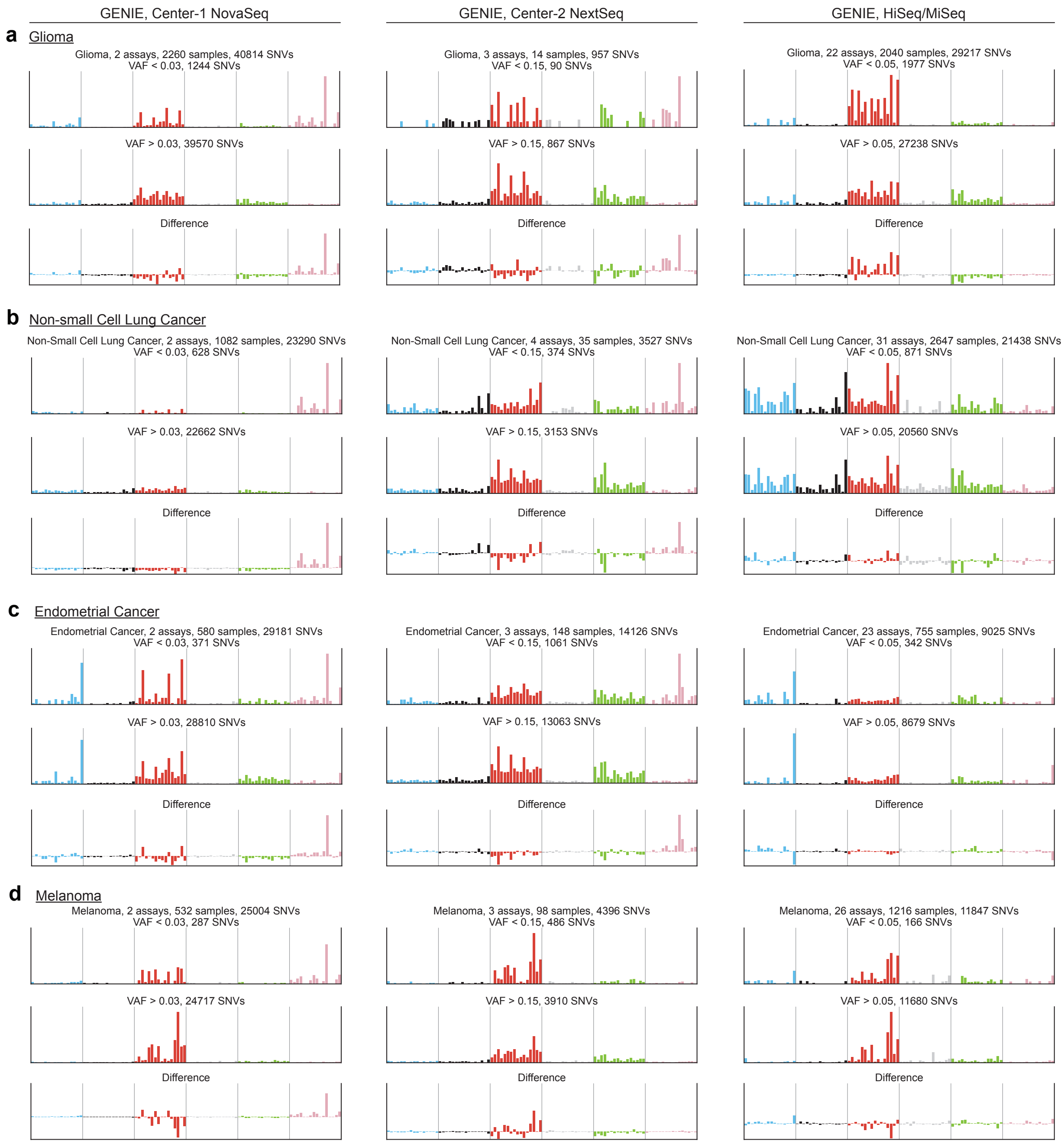


***Supplementary Figure 4. Mutational signature analysis of SNVs in two-color and four-color GENIE samples per tumor type. (a)** Same as Fig. 1d for glioma samples from Center-1-NovaSeq (left), Center-2-NextSeq (middle), and HiSeq/MiSeq assays (right). **(b-d)** Same as (a) for non-small cell lung cancer, endometrial cancer, and melanoma samples, respectively.*

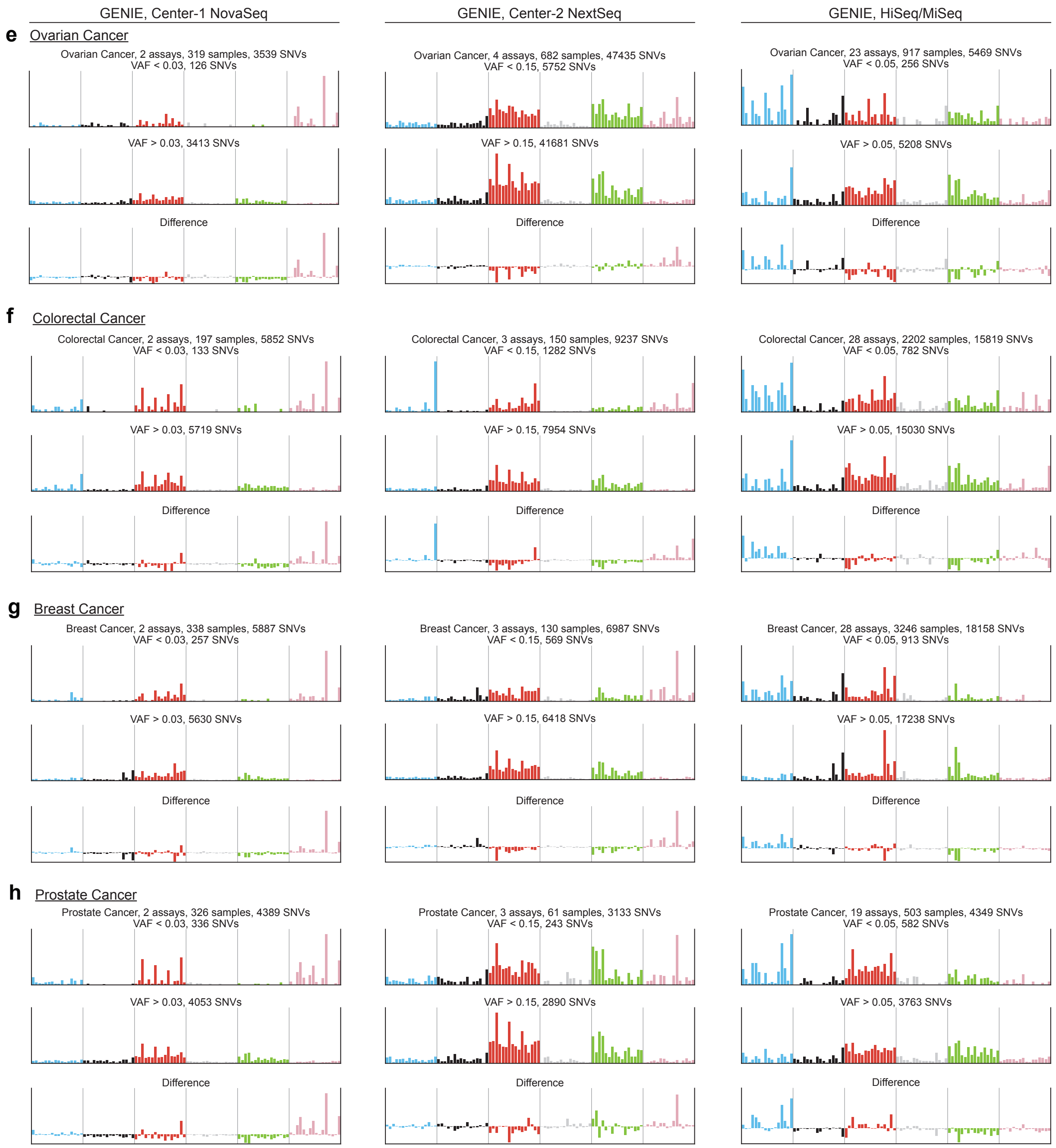


***Supplementary Figure 4 (continued). (e-h)*** *Same as (a) for ovarian cancer, colorectal cancer, breast cancer, and prostate cancer samples, respectively.*

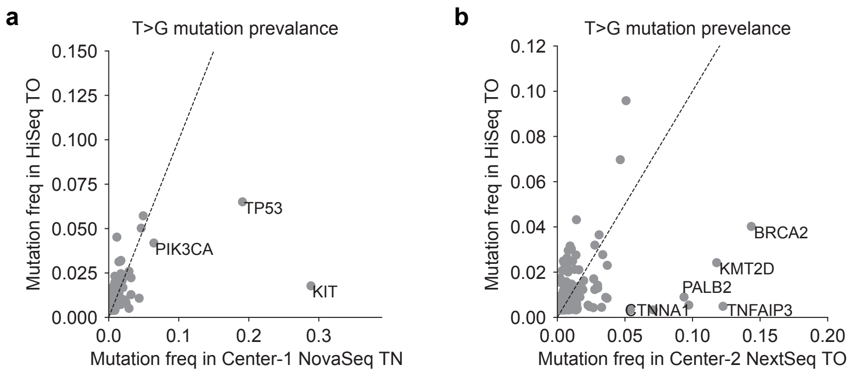


***Supplementary Figure 5. (a)*** *Same as Fig. 2a for Center-1-NovaSeq-TN samples.* ***(b)*** *Same as Fig. 2a for Center-2-NextSeq-TO samples.*

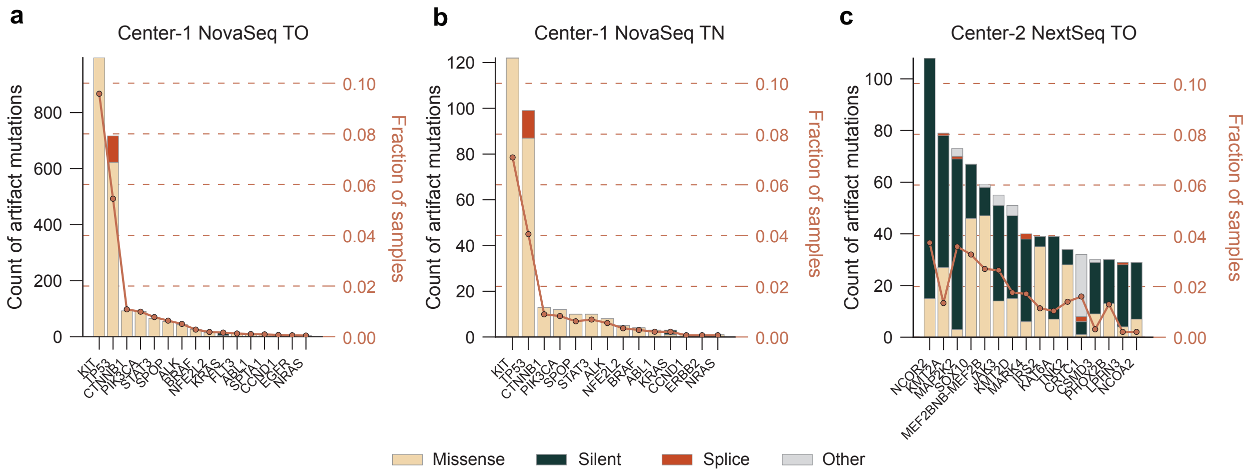


***Supplementary Figure 6.*** *Fraction of samples (dots) and mutation counts (bars) per gene affected by two-color T>G artifact substitutions, stratified by variant classification (missense, silent, splice, other), across three sample sets: Center-1-NovaSeq-TO* ***(a)****, Center-1-NovaSeq-TN* ***(b)****, and Center-2-NextSeq-TO* ***(c)****.*

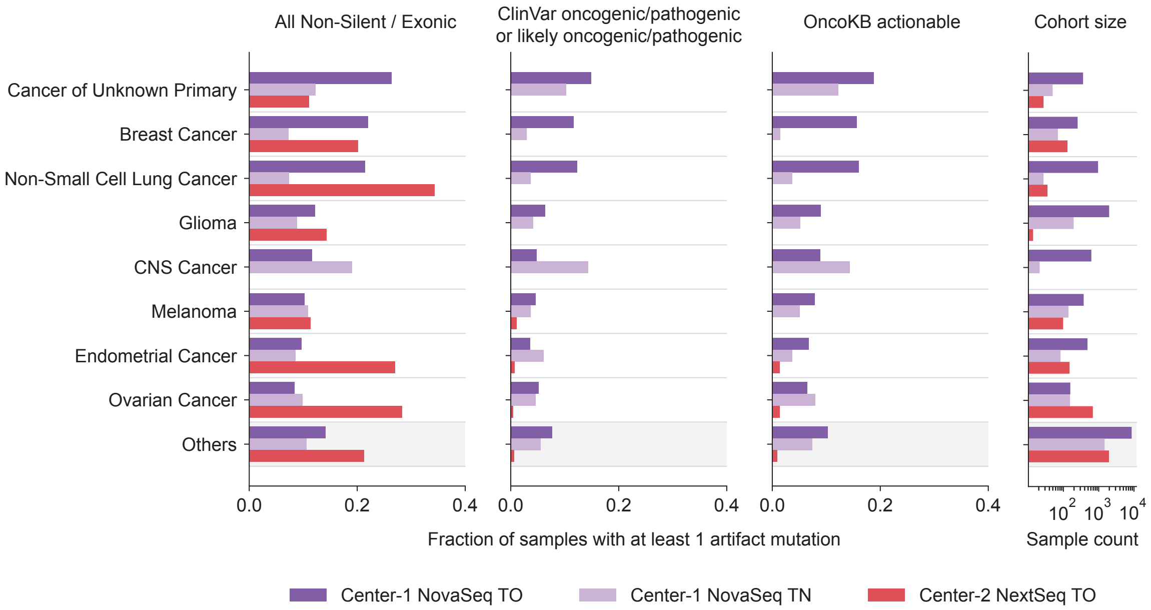


***Supplementary Figure 7. Frequency of two-color T>G artifact mutations across tumor types and clinical variant annotations.*** *Fraction of samples harboring at least one two-color T>G artifact mutation, stratified by tumor type. The top five tumor types with the highest burden are shown for each sample set. Mutations annotated as oncogenic/pathogenic or likely oncogenic/pathogenic (ClinVar) and clinically actionable (OncoKB) are shown separately. Cohort sizes are indicated to the right.*

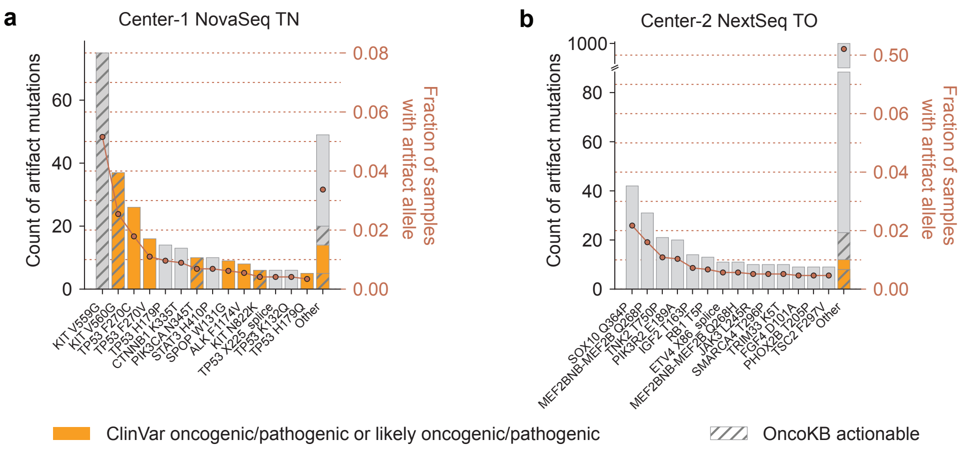


***Supplementary Figure 8.*** ***(a)*** *Same as Fig. 2b for Center-1-NovaSeq-TN samples.* ***(b)*** *Same as Fig. 2b for Center-2-NextSeq-TO samples.*

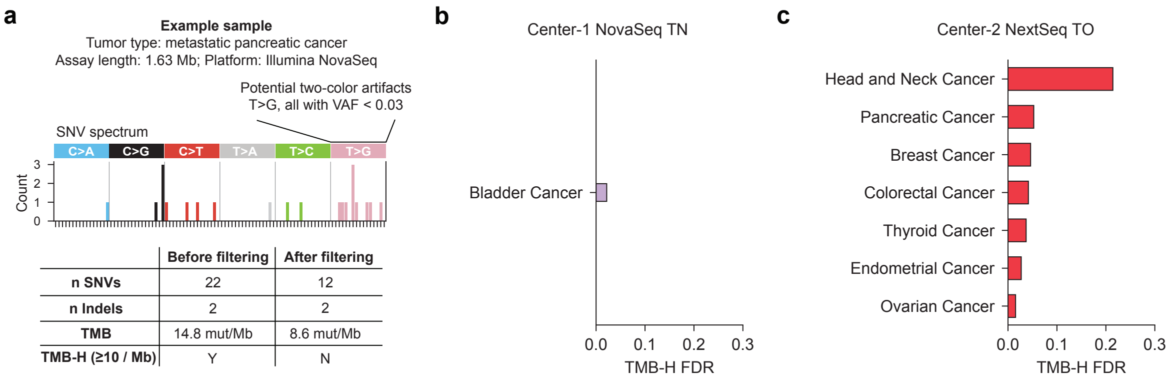


***Supplementary Figure 9. (a)*** *Example showing that two-color T>G artifacts can lead to false-positive identification of TMB-H patients if not removed.* ***(b)*** *Same as Fig. 2c for Center-1-NovaSeq-TN.* ***(c)*** *Same as Fig. 2c for Center-2-NextSeq-TO.*

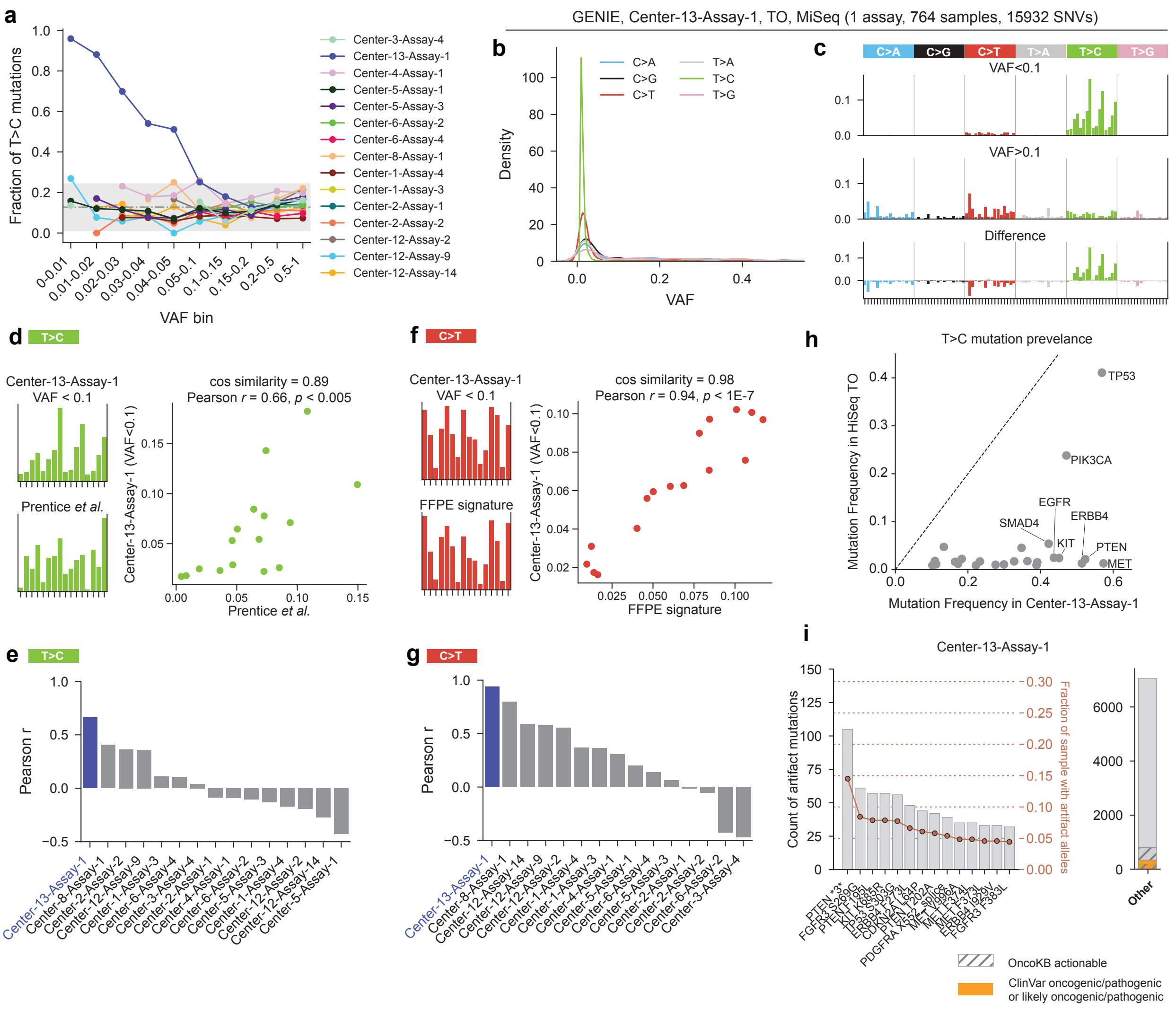


***Supplementary Figure 10. An assay-specific T>C artifact likely caused by insufficient filtering of low-quality variant calls.*** ***(a)*** *Fraction of T>C mutations across VAF bins in different HiSeq/MiSeq assays. Only assays with at least 1,000 SNVs were included.* ***(b-c)*** *Same as Fig. 1c-d for Center-13-Assay-1 samples.* ***(d)*** *Mutational spectrum of low-VAF T>C mutations in Center-13-Assay-1 compared with that of T>C mutations from Prentice et al. For Prentice et al., variant calls from samples in the "fixation group" were included regardless of their quality scores, thus predominantly reflecting low-quality variants.* ***(e)*** *Pearson correlation between the spectrum of low-VAF (VAF<0.1) T>C mutations in different HiSeq/MiSeq assays and that of T>C mutations from Prentice et al.* ***(f)*** *Mutational spectrum of low-VAF C>T mutations in Center-13-Assay-1 compared with that of the C>T FFPE artifact signature from Guo et al.* ***(g)*** *Pearson correlation between the spectrum of low-VAF (VAF<0.1) C>T mutations in different HiSeq/MiSeq assays and that of the C>T FFPE artifact signature from Guo et al.* ***(h)*** *Fraction of samples with a T>C substitution in each gene in Center-13-Assay-1 compared with the HiSeq baseline. Analyses were restricted to the same covered regions, and differences in tumor-type composition across assays were adjusted using a sampling scheme (Methods).* ***(i)*** *Counts of likely T>C artifact alleles (bars) and the fraction of samples carrying each allele (dots) in Center-*

*13-Assay-1. Analyses were restricted to T>C substitutions with VAF<0.1, and only non-silent, non-intronic alleles were included. Variants annotated as oncogenic/pathogenic or likely oncogenic/pathogenic in ClinVar are highlighted in orange. Clinically actionable variants according to OncoKB are indicated by grey hatching. Data from Guo et al. [2] were downloaded from their companion Github repository (https://github.com/QingliGuo/FFPEsig). Data from Prentice et al. [3] were obtained from their Supplementary Tables.*
